# Persistence of rare *Salmonella* Typhi genotypes susceptible to first-line antibiotics in the remote islands of Samoa

**DOI:** 10.1101/2022.03.22.22271657

**Authors:** Michael J. Sikorski, Tracy H. Hazen, Sachin N. Desai, Susana Nimarota-Brown, Siaosi Tupua, Michelle Sialeipata, Savitra Rambocus, Danielle J. Ingle, Sebastian Duchene, Susan A. Ballard, Mary Valcanis, Sara Zufan, Jianguo Ma, Jason W. Sahl, Mailis Maes, Gordon Dougan, Robert E. Thomsen, Roy M. Robins-Browne, Benjamin P. Howden, Take K. Naseri, Myron M. Levine, David A. Rasko

## Abstract

For decades, the remote island nation of Samoa (pop. ~200,000) has faced endemic typhoid fever despite improvements in water quality, sanitation, and economic development. We recently described the epidemiology of typhoid fever in Samoa from 2008-2019 by person, place, and time; however, the local *Salmonella enterica* serovar Typhi (*S.* Typhi) population structure, evolutionary origins, and genomic features remained unknown. Herein, we report whole genome sequence analyses of 306 *S.* Typhi isolates from Samoa collected between 1983 and 2020. Phylogenetics revealed a dominant population of rare genotypes 3.5.4 and 3.5.3, together comprising 292/306 (95.4%) of Samoan versus 2/4934 (0.04%) global *S.* Typhi isolates. Three distinct 3.5.4 genomic sub-lineages were identified and their defining polymorphisms were determined. These dominant Samoan genotypes, which likely emerged in the 1970s, share ancestry with other clade 3.5 isolates from South America, Southeast Asia, and Oceania. Additionally, a 106-kb pHCM2 phenotypically-cryptic plasmid, detected earliest in a 1992 Samoan *S.* Typhi isolate, was identified in 106/306 (34.6%) of Samoan isolates; this is more than double the observed proportion of pHCM2-containing isolates in the global collection. In stark contrast with global *S.* Typhi trends, resistance-conferring polymorphisms were detected in only 15/306 (4.9%) of Samoan *S.* Typhi, indicating overwhelming susceptibility to antibiotics that are no longer effective in most of South and Southeast Asia. This country-level genomic framework can help local health authorities in their ongoing typhoid surveillance and control efforts, as well as to fill a critical knowledge gap in *S.* Typhi genomic data from Oceania.

**IMPORTANCE:** In this study we used whole genome sequencing and comparative genomics analyses to characterize the population structure, evolutionary origins, and genomic features of *S.* Typhi associated with decades of endemic typhoid fever in Samoa. Our analyses of Samoan isolates from 1983 to 2020 identified a rare *S.* Typhi population in Samoa that likely emerged around the early 1970s and evolved into sub-lineages that presently dominate. The dominance and persistence of these endemic genotypes in Samoa are not readily explained by any apparent genomic competitive advantage or widespread acquisition of antimicrobial resistance. These data establish the necessary framework for future genomic surveillance of *S.* Typhi in Samoa for public health benefit.

## INTRODUCTION

Whole genome sequencing (WGS) has become increasingly informative for global surveillance for *Salmonella enterica* serovar Typhi (*S.* Typhi), the causative agent of typhoid fever (1). Following the completion of the first full genome sequence of *S.* Typhi in 2001 (2), WGS has proved integral in exploring *S.* Typhi isolate relationships and relatedness (3), spatial distribution (4), antimicrobial resistance (AMR) (5), virulence (6), and outbreaks (7). Diminishing sequencing costs has permitted large-scale analysis of >1800 *S.* Typhi genomes published in 2015 (8, 9). This facilitated large genomic epidemiology studies of *S.* Typhi collected from endemic countries (10–18) and returning travelers to non-endemic countries (19–23). Genomics has also been used to identify and describe the emergence and international spread of multidrug and extensively drug resistant typhoid fever of great public health concern (11, 24–28), as well as international exchanges of *S.* Typhi (29, 30) primarily among the interconnected hyper-endemic global sub-regions of South Asia, Southeast Asia, and sub-Saharan Africa (31).

Oceania was recently recognized as the fourth global sub-region with a relatively high incidence of typhoid fever (31, 32), while reports spanning multiple decades document the duration of endemic typhoid fever in the region (33–38). Additionally, Oceania has not been a focus of previous large-scale genomic analyses. Excluding Australia and New Zealand, Oceania by nature includes a plethora of small remote island nations scattered across the Pacific Ocean isolated from one another by hundreds to thousands of kilometers of open water. These island nations, especially in the sub-region of Polynesia, are primarily rural, have small capital towns, and generally lack sprawling densely populated metropolitan regions that characteristically sustain high-incidence endemic typhoid through amplified waterborne transmission (39). It has been observed that typhoid fever may behave differently amongst these isolated regions and populations. For example, in a randomized, controlled field trial in Tonga, acetone-inactivated parenteral typhoid vaccine was only modestly protective (34), whereas that same vaccine provided markedly higher efficacy in other field sites, particularly in British Guiana (Guyana) (40), suggesting differences in the host-pathogen relationship by geography.

Samoa, a small island nation (land area ~2,840 km^2^, population ~200,000) located in the Polynesian sub-region of Oceania, has experienced decades of endemic typhoid fever of moderate incidence, despite steady improvements in water supplies and sanitation (41). It is unclear how *S.* Typhi has sustained itself in this remote island population. To explore the population structure and evolutionary origins of *S.* Typhi in Samoa, we characterized the genomic content of isolates collected from 1983 to 2020 in the context of global and ancestral collections of *S.* Typhi. Herein we identify and characterize the dominant *S.* Typhi genotype 3.5.4 and nested genotype 3.5.3, which are exclusive to Samoa and have remained susceptible to antimicrobial agents despite decades of endemicity and liberal prescribing practices. As with other *S.* Typhi sub-lineages we can distinguish these three circulating 3.5.4 sub-lineages by single SNPs. We also estimated the time of emergence of these genotypes to the 1970s and demonstrated their evolutionary relationship to clade 3.5 isolates from Oceania, South America, and Southeast Asia.

## METHODS

This study received ethical clearance from the Health Research Committee of the Ministry of Health (MoH) of Samoa. The University of Maryland, Baltimore (UMB) Institutional Review Board determined this project, protocol HP-00087489, to be exempt.

### Study setting

Samoa is comprised of two main populated islands, Upolu and Savaii. The MoH of Samoa operates two Clinical Microbiology Laboratories, one in Tupua Tamasese Meaole (TTM) Hospital on Upolu and the other in Malietoa Tanumafili II (MT2) Hospital on Savaii. Typhoid fever is a notifiable communicable disease in Samoa that can trigger intense epidemiologic investigation of even individual cases (42, 43). In 2018, the Samoa Typhoid Fever Control Program (TFCP) of the MoH established two Typhoid Fever Epidemiologic “SWAT” Teams (i.e., equipped with specializes epidemiologic tools and tactics), one on each island, trained to perform epidemiological investigations of the household (and/or workplace or school) of every bacteriologically confirmed case of typhoid fever occurring anywhere in Samoa. The Typhoid SWAT teams ascertain the clinical status of all contacts in the case household (or workplace, or school), obtain stool cultures from contacts to detect asymptomatic excreters of *S.* Typhi, assess environmental risk factors for typhoid transmission, and seek evidence of links to other typhoid cases.

### Bacterial isolates sequenced in this study

This study includes contemporary *S.* Typhi isolates collected in Samoa by the MoH Clinical Microbiology Laboratories and from the Typhoid Fever Public Health Laboratory (from stool cultures obtained by the Typhoid SWAT Teams). It also includes historical *S.* Typhi isolates collected from travelers who acquired typhoid in Samoa (but were diagnosed upon return to their home country); these travel-associated isolates were either made available for sequencing by collaborators or had been previously characterized (Tables S1 and S2).

From April 2018 through June 2020, *S.* Typhi isolates from blood cultures of acutely febrile patients were bacteriologically confirmed by standard methods (41, 44–46), tested for antimicrobial susceptibility (AST), and stored at −70°C. Cases of typhoid fever refer to patients with clinical signs and symptoms of enteric fever (47) that triggered the collection of a blood culture from which *S.* Typhi was isolated and confirmed. Under the routine public health surveillance activities of the MoH of Samoa, every case of typhoid fever was rapidly reported to one of the MoH Typhoid SWAT Teams. During household (or other venue) visits, up to three rectal swabs were collected ~1-2 days apart from asymptomatic contacts, transported in Cary-Blair transport media to the Typhoid Fever Public Health Laboratory and examined using standard bacteriology methods (44, 45). These asymptomatic, subclinical *S.* Typhi infections detected by stool culture in household contacts of a confirmed acute clinical case are referred to as asymptomatic shedders.

All stored *S.* Typhi isolates from cases and asymptomatic shedders were shipped from Samoa to the Microbiological Diagnostic Unit Public Health Laboratory (MDU PHL) at The Peter Doherty Institute for Infection and Immunity in Melbourne, Australia for whole genome sequencing (Table S1). Additionally, from the MDU PHL historical freezer collection, all 14 previously uncharacterized historical isolates of *S.* Typhi originating from travelers from Samoa spanning the years 1983-2011 were included in this study (Table S1).

### Phenotypic antimicrobial susceptibility testing (AST)

At the TTM and MT2 Clinical Microbiology Laboratories, *S.* Typhi blood isolates from 2018 to 2020 were tested for phenotypic AST by the Kirby-Bauer Disc Diffusion method (48) following the 2009 Clinical and Laboratory Standards Institute (CLSI) breakpoints (49) until 2019, when a switch was made to the more cost-effective European Committee for Antimicrobial Susceptibility Testing (EUCAST) guidelines (50).

### DNA extraction and whole genome sequencing (WGS)

DNA extraction and WGS were performed at the MDU PHL following standard procedures (51). Unique dual-indexed libraries were prepared using the Nextera XT DNA sample preparation kit (Illumina). Libraries were sequenced on the Illumina NextSeq 500 with 150-cycle paired-end chemistry. Illumina sequence reads underwent preliminary inspection using the MDU PHL bioinformatics pipeline, Nullarbor (52).

### Draft genome assembly

Illumina reads were quality filtered and trimmed using Trimmomatic v0.38 (53) and assembled *de novo* using SPAdes v3.14.1 (54) in Python v2.7.14. The final draft assemblies were filtered to contain only contigs ≥500 bp in length and with ≥5x k-mer coverage, as previously described (55). One isolate (AUSMDU00049311) not meeting quality metrics was excluded from subsequent analyses.

### Read alignment and genotyping

Genotypes were assigned according to GenoTyphi (9, 56). Paired-end Illumina reads were mapped to the reference strain CT18 (GenBank Accession AL513382.1), using Snippy v4.6.0 (57). GenoTyphi v1.9.1 was then used to analyze the Snippy output vcf files. GenoTyphi uses a set of core genome SNPs relative to CT18 to sequentially classify *S.* Typhi genomes into primary clusters/clades, clades, subclades, and sub-lineages represented by a numerical sequence of one, two, three, or four digits, respectively.

### Complete genome sequencing and assembly

Fifteen Samoan *S.* Typhi strains were also selected to undergo long-read sequencing to obtain a complete genome. Within each genotype of *S.* Typhi presently circulating in Samoa, the oldest and most recent isolates were selected for genome closure at the MDU PHL using established pipelines (58, 59). Long-read sequencing libraries were prepared using the Oxford Nanopore Technologies ligation sequencing kit (SQK-LSK109) and sequenced on the GridION system using R9.4.1 flow cells. Final closed/completed assemblies were polished using the full long-read set with Medaka v1.0.3 (60) and with the short-read set using Pilon v1.23 (61).

### *S.* Typhi genomes from other studies

High-throughput short-read Illumina data for all Samoan *S.* Typhi genomes (n=117) and all non-Samoan clade 3.5 genomes (n=12) previously published by Wong et al. (8, 9) were downloaded from the European Nucleotide Archive (ENA) BioProject PRJEB3215. Duplicate Samoan isolates were removed. Illumina reads for all clade 3.5 genomes from Colombia (n=20) and Chile (n=68) were downloaded from ENA BioProjects PRJEB42858 (62) and PRJEB20778 (63), respectively. These *S.* Typhi sequences from other studies and their accession numbers are listed in Table S2. All short read Illumina data from these studies were assembled using SPAdes v3.14.1 (54), as described above.

### Global collection and *in silico* typing

Pathogenwatch is an online bacterial genome surveillance tool for *S.* Typhi (64, 65) with *in silico* typing modules for uploaded data and a global collection of published *S.* Typhi assemblies for comparison (65). All genome assemblies from this study (Tables S1-S3) were uploaded to Pathogenwatch for plasmid replicons detection using the PlasmidFinder (66) Enterobacteriaceae database implemented as “IncTyper” in Pathogenwatch. Genomic predictions of AMR were determined using the Pathogenwatch curated AMR library, which includes both genes and point mutations known to confer phenotypic resistances (64).

### Maximum-likelihood phylogenies

A global phylogeny based on the single nucleotide polymorphisms (SNPs) of the core sequences of assembled genomes was generated following similar configurations to Wong et al. (9). For all assemblies in this study and global assemblies downloaded from Pathogenwatch, SNPs were determined against the reference strain CT18 using NASP v1.2.0 (67). To exclude regions of recombination, the resultant alignment was analyzed with Gubbins v2.4.1 (68). RAxML v8.2.10 (69) was run on the PHYLIP format alignment of filtered polymorphic sites using the generalized time-reversible (GTR) site substitution model with discrete Gamma (Γ) distributed rate variation and the Lewis ascertainment bias correction (ASC_GTRGAMMA), and 100 bootstrap pseudo-replicates. The resulting phylogeny was rooted at genotype 0.0 and visualized in iTOL (70).

For a clade-specific phylogeny, these steps were repeated with clade 3.5 assembled genomes aligned to a local 2012 Samoan *S.* Typhi reference strain H12ESR00394-001A (GenBank Accession LT904890.1). Pairwise SNP distances between genomes were calculated using snp-dists v0.8.2 (71). The resulting phylogeny was midpoint rooted and visualized in iTOL (70). This was repeated with strain CT18, and unique SNPs defining the Samoan 3.5.4 sub-lineages were manually identified from global and clade 3.5 NASP matrices using the criteria established in GenoTyphi for the identification and characterization of canonical SNPs (9).

### Whole genome and pHCM2 gene content analyses

To assess whether any unique or lost genes may impart a competitive advantage to the Samoa dominant genotypes, a Large Scale BLAST Score Ratio (LS-BSR) analysis (72, 73) was performed on draft and complete genomes (Tables S1-S2) including a diverse collection of complete reference genomes from GenBank spanning 1916 to 2019, derived from typhoid endemic regions, and representing most major *S.* Typhi phylogenetic groups (Table S3). The LS-BSR output matrix (Table S4) encodes a measure of homology of all predicted protein-coding sequences (genes) across all genomes in the comparison to examine total gene content. As the *S.* Typhi organism is genomically conserved and monophyletic (74, 75), a stringent threshold to define a putative gene as present was set to BSR ≥0.9. Less than 0.9 was considered absent. Scoary v1.6.16 (76) was then used to examine this matrix against multiple variables (Table S5). Genes with a p-value <0.05 adjusted with Bonferroni’s adjustment method for multiple comparisons were considered significant. The predicted protein function of each gene was determined using an in-house annotation pipeline (77).

To confirm the presence/absence of the full-length pHCM2 identified by the IncFIB(pHCM2) replicon in Pathogenwatch, LS-BSR analysis was completed on all Samoan *S.* Typhi draft genomes using the 106,710 bp pHCM2 molecule from the complete genome of the 1992 Samoan strain AUSMDU00051359 (GenBank Accession CP090227; Table S1) annotated in RAST (78–80) as input (Table S7). To assess homology of the available complete pHCM2 molecules, all four circularized pHCM2-like molecules present in the completed Samoan *S.* Typhi genomes (this study) were compared against all five pHCM2-like molecules present in the GenBank reference collection.

### Temporal and evolutionary analyses

For temporal and evolutionary phylogenetic analyses, raw reads for all clade 3.5 isolates were first aligned to the local Samoan reference strain H12ESR00394-001A and SNPs were determined using Snippy v4.6.0 (57), with filtering of phage regions identified using PHASTER (81). The final core SNPs were extracted using Snippy core v4.6.0 (57) and used to construct a maximum-likelihood (ML) phylogeny in MEGAX v10.1.8 (82, 83) using the GTR+Γ substitution model and 100 bootstrap pseudo-replications. The resulting NEWICK tree was examined in TempEst v1.5.3 (84). Tree tips were assigned their sampling time and a regression analysis of the root-to-tip branch distances as a function of sampling times was conducted with the best-fitting root selected according to heuristical residual mean squared, similar to previous studies (13, 85).

To estimate a date of divergence of a most recent common ancestor (MRCA), the same core alignment was analyzed in BEAST v1.10.4 (86). The molecular clock model was calibrated using dates of isolation and specified with the GTR+Γ nucleotide substitution model, uncorrelated relaxed (UCR) clock model, and exponential growth coalescent tree priors. The number of constant nucleotide patterns were specified as the number of specific nucleotides. Three independent runs performed using a Markov chain Monte Carlo length of 200 million steps, sampling every 20,000 steps, were then combined using LogCombiner v1.10.4 (87) and assessed in Tracer v1.7.1 (88) to verify that the effective sample size (EES) of all key parameters was at least 200. The resultant summary maximum clade credibility (MCC) tree was constructed using TreeAnnotator v1.10.4 (87) after discarding the first 10% of iterations as burn-in and using median node-height values. The MCC tree was decorated in R v4.1.1 using the ggtree package (89–91). All above steps were also repeated using the classical reference genome for *S.* Typhi strain CT18.

### Data availability

All raw sequences and complete assemblies generated in this study have been deposited on GenBank BioProject PRJNA319593 with the accessions listed in Table S1. All assemblies analyzed have been deposited on Figshare (doi: 10.6084/m9.figshare.18665686).

## RESULTS

### Number and sources of Samoan *S*. Typhi sequences

During the study period of April 2018 to June 2021, the MoH of Samoa stored and shipped 173 blood isolates from culture-confirmed acute cases of typhoid fever and 15 rectal swab isolates from asymptomatic shedders of *S.* Typhi (Table S1). One 2020 Samoan sequence (AUSMDU00049311) from an acute case was removed due to poor assembly metrics. Together with the 14 Samoan *S.* Typhi isolates from the MDU PHL historical freezer collections sequenced in this study (Table S1) and the 105 non-duplicate Samoan *S.* Typhi previously published by Wong et al. (8) (Table S2), we examined a final collection of 306 Samoan *S.* Typhi. Most isolates were from 2012 and 2018 to 2020 (Fig. 1).

**Fig. 1.**
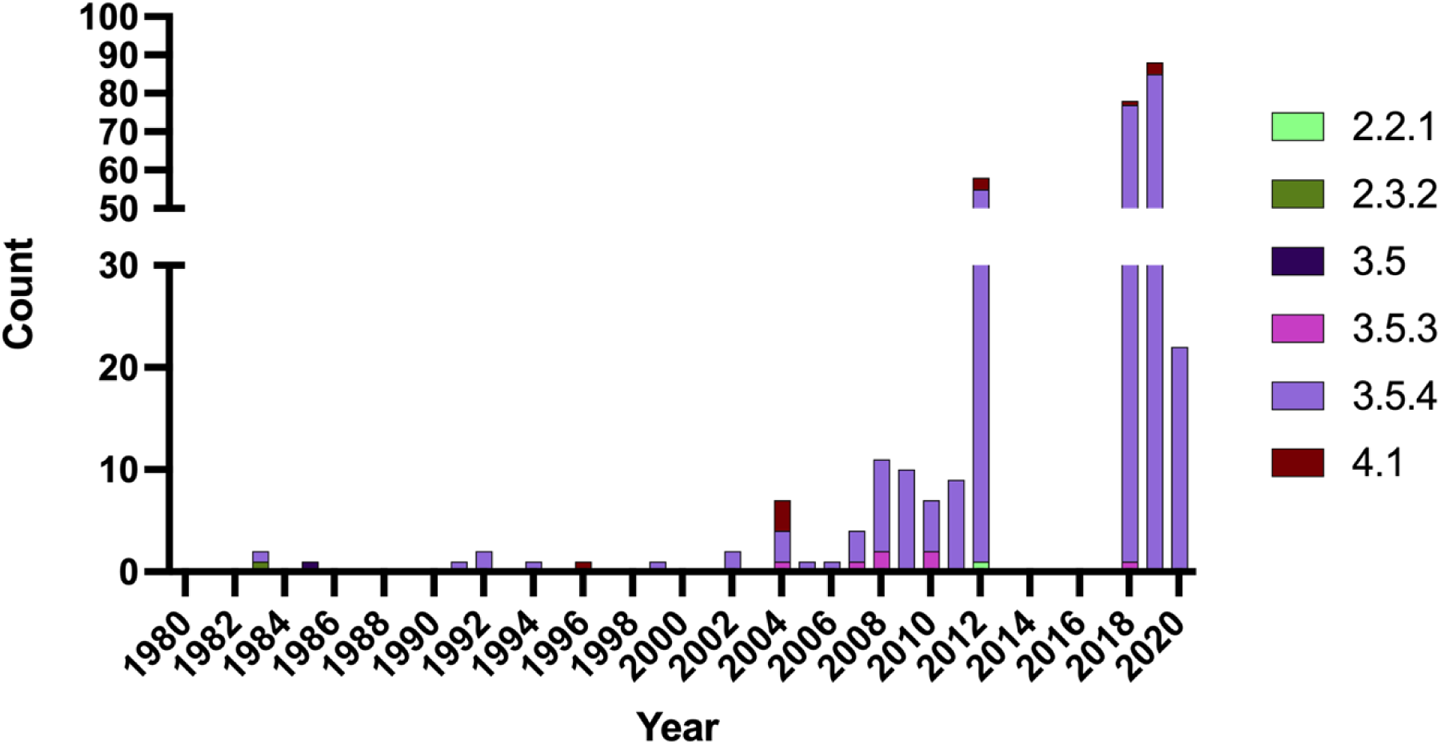
Frequency and genotype distribution of Samoan *S.* Typhi sequenced isolates by year of collection. Samoan isolates of *S.* Typhi (N=306) analyzed in this study span the years 1983-2020. Isolates from 1983-2011 were all collected from travelers from Samoa in Australia and New Zealand. Most isolates in 2012 and all isolates in 2018-2020 were collected in Samoa by the Ministry of Health. Genotype designations were determined using GenoTyphi (9, 56) on paired-end Illumina reads mapped to strain CT18.

### Phylogenetic structure of *S.* Typhi from Samoa

The population structure of *S.* Typhi from Samoa from 1983 to 2020 was dominated by isolates in parent clade 3.5 (Fig. 2A), namely genotypes 3.5.4 (285/306; 93.1%) and 3.5.3 (7/306; 2.3%). Genotypes 3.5.4/3.5.3 appeared to be exclusively associated with Samoa as they have not yet been reported elsewhere in the world, apart from two travel-related isolations of unknown origin recovered in Australia in 2011 (8) (Table S2). Within the Samoa-exclusive genotype 3.5.4, we identified three distinct sub-lineages numbered 1-3 (genotype designations 3.5.4.1, 3.5.4.2, and 3.5.4.3) (Fig. 2B). These sub-lineages can be exclusively delineated from other *S.* Typhi by the SNPs listed in Table 1.

**Fig. 2.**
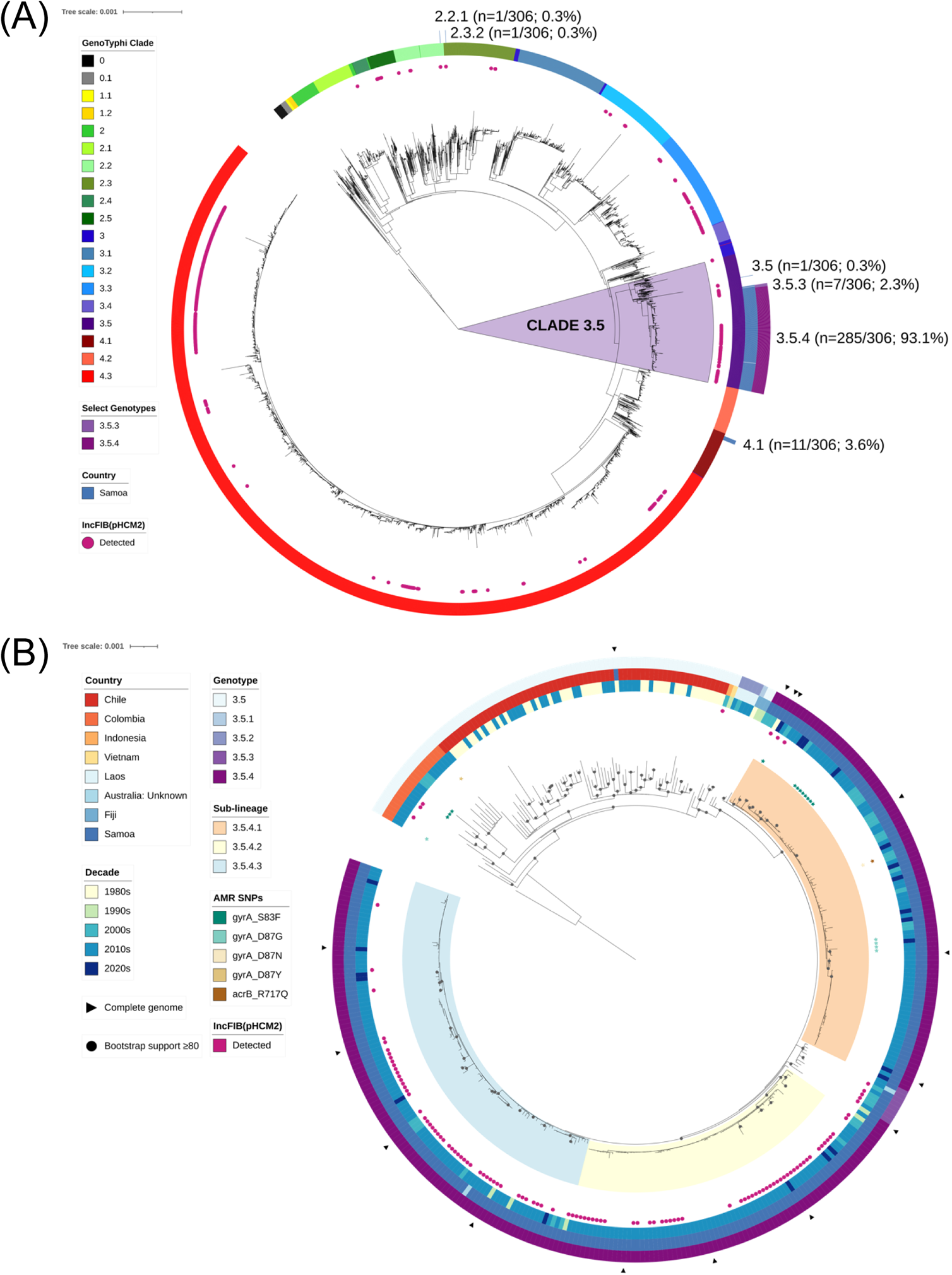
Phylogenetic analysis of Samoan *S.* Typhi genomes in a global and dominant clade-specific context. **(A)** Maximum-likelihood tree of 5,240 *S.* Typhi genome assemblies using reference strain CT18 (GenBank Accession AL513382.1) including 306 from Samoa with GenoTyphi clade (ring 1) [innermost ring] (9, 56), country (ring 2) [Samoa], and genotypes 3.5.4 and 3.5.3 (ring 3) labeled. The distribution of the IncFIB(pHCM2) replicon detected using with PlasmidFinder (66) is indicated with pink shaded circles. Clade 3.5 genomes are highlighted with a light purple background. **(B)** Maximum-likelihood phylogeny of assembled clade 3.5 genomes (N=394) using a local Samoan reference strain H12ESR00394-001 (GenBank Accession LT904890.1). Country of origin, decade of isolation, antimicrobial resistance point mutations and the detection of the IncFIB(pHCM2) replicon are displayed.

**Table 1.** Candidate canonical SNPs delineating Samoan genotype 3.5.4 sub-lineages

| Genotype | n/N* | Ref.<br>allele | Alt.<br>allele | CT18<br>position | Gene | Protein |
| --- | --- | --- | --- | --- | --- | --- |
| 3.5.4.1 | 96/96 | G | A<sup>2</sup> | 3960783 | STY4102 | putative L-lactate dehydrogenase |
|  |  | G | A<sup>2</sup> | 4794120 | *gpmB* | probable phosphoglycerate mutase 2 |
| 3.5.4.2 | 77/77 | C | T<sup>2</sup> | 1731534 | STY1812 | exodeoxyribonuclease III |
|  |  | C | T<sup>2</sup> | 1945934 | STY2094 | glucose 6-phosphate dehydrogenase |
| 3.5.4.3 | 106/107<sup>1</sup> | C | T<sup>3</sup> | 295362 | STY0278 | putative drug efflux protein |

Genotype = Suggested sub-lineage designation; n/N = number of *S. Typhi* isolates in Samoa over total number of *S. Typhi* isolates in the sub-lineage; Ref. Allele = CT18 allele; Alt. allele = alternative SNP candidates in the genotype sub-lineage.
\*All SNPs were detected in 100% of their designated genotype groupings (N) and absent from 100% of out-grouped isolates (5,240 global *S. Typhi* genomes were analyzed)
<sup>1</sup> One 3.5.4.3 isolate is of unknown origin.
<sup>2</sup> Synonymous intragenic silent mutation
<sup>3</sup> Non-synonymous intragenic missense mutation

A small cluster of Samoan *S.* Typhi isolated from 1996 to 2020 were genotype 4.1 (11/306; 3.6%), and single isolates of genotypes 2.2.1, 2.3.2, and 3.5 were detected in 2012, 1983, and 1985, respectively (Fig. 2A). During the period of intensified typhoid surveillance, i.e., 2018 to 2020, only genotypes 3.5.3, 3.5.4, and 4.1 were identified as circulating in Samoa (Fig. 1).

Clade 3.5 (i.e., genotypes 3.5, 3.5.1, 3.5.2, 3.5.3, and 3.5.4) includes isolates sampled over the past 40 years from Samoa (n=293), Chile (n=68), Colombia (n=20), Laos (n=5), Fiji (n=3), Indonesia (n=1), Vietnam (n=1) and Australia from unknown travel (n=2) (Tables S1-S2). A ML phylogeny inferred from all clade 3.5 genomes demonstrated that the Samoan genotypes 3.5.4 and 3.5.3 do not intermingle with isolates from these other countries, despite sharing a common core genome indicating shared evolutionary ancestry (Fig. 2B). The only Samoan isolate of genotype 3.5 (AUSMDU00021621) was isolated in 1983 and is phylogenetically most similar to isolates from Chile in the 1980s (Fig. 2B). Collectively, these data demonstrate that the genotypes 3.5.4 and 3.5.3 are exclusive to Samoa and have been present for decades with only sporadic isolations of alternative genotypes.

### Clock-like behavior and date of emergence of common ancestor to Samoa-exclusive *S.* Typhi genotypes

Root-to-tip regression analysis as a function of sampling time was performed on the clade 3.5 ML tree using TempEst (84) with the best-fitting root selected (Fig. 3A). We observed a positive temporal association between year isolated and root-to-tip evolutionary distance, with an R^2^ value of 0.302. This regression coefficient is comparable to those reported in other studies of *S.* Typhi (13, 29, 85).

**Fig. 3.**
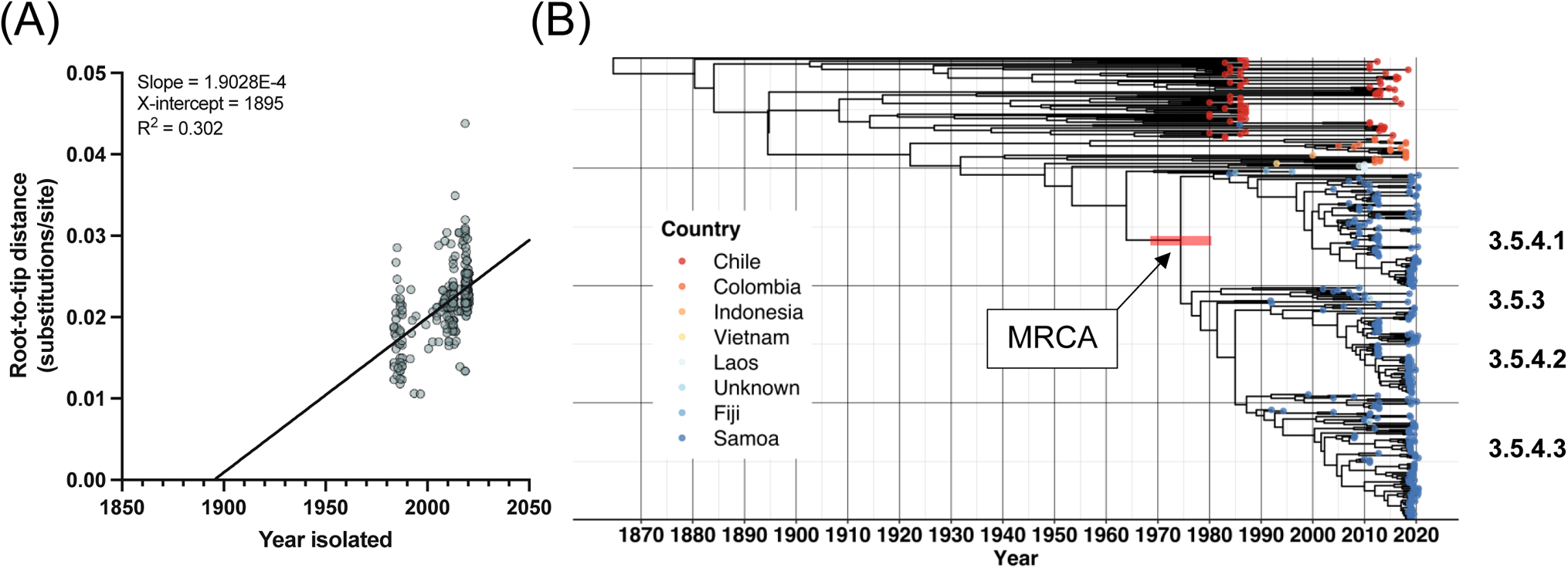
Temporal and evolutionary analysis of *S.* Typhi clade 3.5. **(A)** Temporal signal analysis of clade 3.5 *S.* Typhi. TempEst v1.5.3 (84) root-to-tip regression analysis as a function of sampling time is shown for the core sequence alignment against the local Samoan reference strain H12ESR00394-001A (GenBank Accession LT904890.1) of all 394 *S.* Typhi belonging to clade 3.5 (isolated between 1983 and 2020 with variable date precision) with the best-fitting root selected according to heuristical residual mean squared. **(B)** Bayesian evolutionary analysis of the same core sequence alignment showing the maximum-clade credibility (MCC) phylogenetic tree of Samoan genotypes 3.5.4 and 3.5.3 isolates in the context of global clade 3.5 *S.* Typhi isolates. The age of the node of the most recent common ancestor (MRCA) to the Samoan genotypes 3.5.3 and 3.5.4 isolates (arrow) had a mean of 1974 and the red horizontal bar represents the 95% Highest Posterior Density (HPD: 1968, 1981) of the node. Tip colors indicate country of origin for each isolate. A time scale is shown along the x-axis is relative to the most recent sampling date of 2020-06-09. The genotype/sub-lineage designations are labeled to the right. The tree was constructed with BEAST v1.10.4 (87) and visualized using the ggtree package in R (89–91).

To infer a date of emergence of the MRCA of genotypes 3.5.4 and 3.5.3, we analyzed the summary statistics and MCC tree output of our clade 3.5 BEAST analysis (87) (Fig. 2B). The date of emergence of the MRCA to the dominant Samoa genotypes (3.5.4 and 3.5.3) was estimated to be around 1974 (95% Highest Posterior Density [HPD]: 1968, 1980). The substitution rate (meanRate parameter in BEAST) was 5.82×10^−8^ (95% HPD: 5.0×10^−8^, 6.6×10^−8^) genome-wide substitutions per site per year, or ~0.27 (95% HPD: 0.24, 0.32) substitutions per genome per year. Compared with a rate of ~0.8 SNPs per genome per year calculated for genotype 4.3.1 (13, 85) (previously designated the H58 haplotype (8)), these estimates represent a reduced rate of SNP acquisition across the core genome in the dominant Samoan genotypes compared to genotypes in other geographic locations.

### Unique gene-content analysis

To determine if any genetic characteristics might explain the persistence and endemic nature of the *S*. Typhi in Samoa, an all-versus-all gene-by-gene comparative analysis was performed using LS-BSR (72, 73) including all Samoan and clade 3.5 isolates (Tables S1-2) versus a diverse collection of 47 complete isolates from GenBank (Table S3). The number and annotation of significantly differential genes (uniquely present or absent with p<0.05 adjusted with Bonferroni’s method for multiple comparisons correction) were identified based on metadata groupings. There were >500 differential genes among *S.* Typhi of Samoan origin versus non-Samoan isolates, as well as in the Samoan genotypes 3.5.4/3.5.3 versus other genotypes. Each 3.5.4 sub-lineage contained between 200-270 differential genes distinguishing them from the other sub-lineages and genotypes. With respect to time, ~200 differential genes were encoded in *S.* Typhi from the 1980s compared to only <15 across more recent decades. These differential genes by each trait are detailed in Table S5. Within each trait, roughly 30-60% of the significant differential genes were hypothetical or putative in their functional annotation, which is not uncommon. Of the genes uniquely present in Samoa or genotypes 3.5.4/3.5.3, which are most descriptive of Samoan *S.* Typhi, the functional annotations predominantly included regulatory genes, metabolic factors, and mobile elements (Table S5). No genes known to confer enhanced virulence or antimicrobial resistance to first-line antibiotics were detected.

### Antimicrobial resistance

Among the Samoan *S.* Typhi isolates, 15/306 (4.9%) contained a point mutation in DNA gyrase subunit A, a quinolone resistance determining region (QRDR) conferring predicted intermediate resistance to fluoroquinolones (64), which are commonly prescribed in the treatment of typhoid fever (47) (Fig. 1B, Table S6). These were *gyrA*-S83F (n=10/15), *gyrA*-D87G (n=4/15), and *gyrA*-D87N (n=1/15). Of these 15 isolates with predicted resistance, ten were isolated in Samoa in the Clinical Microbiology Laboratories and phenotypic AST assays for ciprofloxacin were completed and available for eight of ten isolates. All eight isolates were reported susceptible to ciprofloxacin (Table S6) indicating disagreement with the predicted intermediate resistance and the genotype. Azithromycin is an oral macrolide drug also commonly prescribed in the treatment of typhoid fever (47). An additional point mutation in acriflavin resistance protein B gene, *acrB*-R717Q, known to limit susceptibility to azithromycin (15), was also identified in the isolate containing the *gyrA*-D87N point mutation. Unfortunately, we did not have access to this isolate to perform AST because it was collected in New Zealand from a traveler from Samoa.

### Comparative analysis of pHCM2 plasmid

Using IncTyper in Pathogenwatch (64), the plasmid replicon corresponding to IncFIB(pHCM2) was detected in 106/306 (34.6%) of Samoan *S.* Typhi isolates (Figs. 1B and 4A). This replicon is significantly (p<0.01, chi-square test of independence) associated with *S.* Typhi from Samoa, compared with 618/4934 (12.5%) of non-Samoan global *S.* Typhi. The pHCM2 sequence from the 1994 Samoan isolate, AUSMDU00051359, was completed and circularized and is 106,710 bp in length (GenBank Accession CP090228; Table S1). The plasmid contains one tRNA gene sequence and 129 genes, of which 89/130 (68.5%) encode a hypothetical protein with no known function (Table S7). The remaining 41 genes include functions for replication, central metabolism or mobile elements. No genes for antimicrobial resistance or virulence were detected. The full-length pHCM2 was confirmed in three other complete Samoan genome sequences from 2008, 2019, and 2020 selected for closure (Table S1). All four completed circularized plasmids were identical in length (Table S1) and gene content (Fig. 4), despite more than 25 years between the oldest and most recent isolation.

**Fig. 4.**
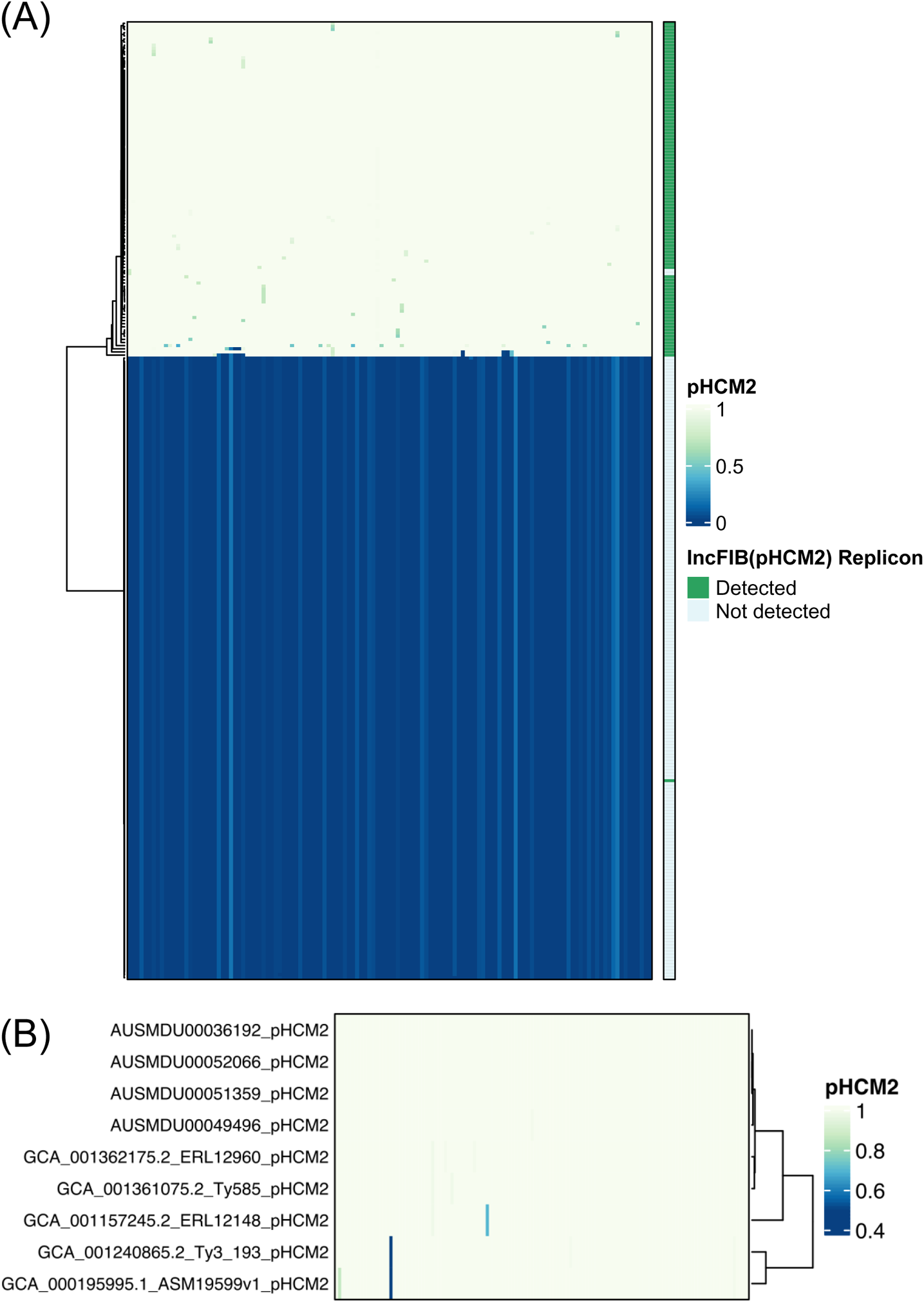
LS-BSR heatmap identifying pHCM2 genes in Samoan *S.* Typhi. **(A)** The 130 sequential coding regions in the circularized complete pHCM2 plasmid of 1994 Samoan strain AUSMDU00051359 was compared all Samoan *S.* Typhi (n=306) draft genomes for gene homology. The heatmap depicts gene absence (dark blue) and presence (light yellow) as a BLAST score ratio from 0 to 1, respectively. The bar on the right indicates the presence (green) or absence (light blue) of the IncFIB(pHCM2) replicon as detected by IncTyper (64). **(B)** This comparison was repeated for all four complete circularized pHCM2 molecules in Samoan *S.* Typhi and the five GenBank complete reference genomes that contain a complete circularized pHCM2 molecule. Sequential gene homology compared with the pHCM2 molecule from strain AUSMDU00051359 is depicted, with homology ranging from low (dark blue) to high (light yellow). Strain ERL12960 is genotype 4.3.1.1 isolated in India in 2012. Strain Ty585 is genotype 4.3.1.1 isolated in Bangladesh in 2012. Strain ERL 12148 is genotype 4.3.1.1 isolated in India in 2012. Strain Ty3 is genotype 4.3.1.1 isolated in Vietnam in 1997. Strain ASM19599v1 is strain CT18, a genotype 3.2.1 isolate from Vietnam in 1993. The GenBank accession numbers are listed in Table S3.

A focused LS-BSR analysis of the 130 pHCM2 genes against all 306 Samoan *S.* Typhi strains (Fig. 4A) revealed the presence of the pHCM2 genes in 106/306 (34.6%) of the isolates. In two isolates, 10071_8_49_Sam0049_2012 and AUSMDU00049494, LS-BSR detected the majority of the gene content of the pHCM2, but the >90% coverage of the replicon region required for IncTyper (64, 65) was lacking from the draft assemblies, resulting in omission from identification via incompatibility typing. In contrast, in the 1983 Samoan isolate AUSMDU00021620, IncTyper identified the IncFIB replicon, but the pHCM2 gene content was absent from the analyzed assembly. The remaining replicon determination resulted in an agreement of the replicon and the presence of remainder of the pHCM2 gene content.

Comparison of plasmid gene content between the four complete Samoan pHCM2 molecules and five GenBank complete reference sequences identified a near complete plasmid homology (Fig. 4B). Only two hypothetical genes, 220341.113.peg.18 and 220341.113.peg.2 (Table S7) demonstrated divergence in pHCM2 from strain CT18 compared with the pHCM2 molecules of four Samoan strains, indicating a significant level of cryptic plasmid conservation.

## DISCUSSION

The phylogenetic analyses in this study demonstrate that 95.4% (292/306) of *S.* Typhi isolates from Samoa, spanning 1983 to 2020, are of rare genotypes 3.5.4 and 3.5.3 (Fig. 1–2, Table S1). Among non-Samoan *S.* Typhi, genotypes 3.5.4 and 3.5.3 were recorded through passive surveillance in only two Australian isolates of unknown travel origin (Fig. 2A). Considering that 41/306 (13.4%) of the Samoan *S.* Typhi were collected in Australia from travelers to Samoa (Tables S1 and S2) and the frequent migration and travel between Samoans and their extended families living in Australia, it is plausible that these two unknown isolates of genotypes 3.5.4 and 3.5.3 may have been acquired in Samoa. Additionally, the intermingling of these imported isolates of unknown origin amongst the Samoan *S.* Typhi further supports this putative association (Fig. 2B).

Therefore, we consider the genotypes 3.5.4 and 3.5.3 as globally exclusive to Samoa. Within these dominant genotypes, we defined three discrete genotype 3.5.4 sub-lineages circulating in Samoa via core genomic variation and canonical SNPs (Fig. 2B; Table 1). These sub-lineages represent distinct core genome evolution detectable by comparative genomic analysis. Including the relatively infrequent genotypes 3.5.3 and 4.1 in this Samoan *S.* Typhi collection, a total of five genotypes/sub-lineages of Samoan *S.* Typhi distinguishable by single SNPs form a functional WGS framework with public health utility. Specifically, future isolates may be sequenced, rapidly categorized by genotype/sub-lineage, and utilized to infer putative linkages with other genomically related isolates for targeted investigation by the Samoan Typhoid SWAT teams (92).

Phylogenetic ancestry of genotypes 3.5.4/3.5.3 is shared with clade 3.5 *S.* Typhi isolates from countries in Oceania, South America, and Southeast Asia (Fig. 2B). Genotypes 3.5.4/3.5.3 are predicted to have emerged from the clade 3.5 genomic background in ~1974 (95% HPD: 1968-1980) (Fig. 3B). However, *S.* Typhi has been present in Samoa decades prior. Endemic typhoid fever in Samoa was recorded in World Health Organization surveillance data in the 1950s (93) and in epidemiological modeling data collected in the 1960s (33). These records corroborate a persistently endemic situation predating the estimated emergence of genotypes 3.5.4/3.5.3. It is not known whether *S.* Typhi in Samoa in the 1950s and 1960s were predecessors of the 3.5.4/3.5.3 Samoan *S.* Typhi or other genotypes that were eventually displaced by the 3.5.4/3.5.3 Samoan *S.* Typhi.

The estimated core genome substitution rate of clade 3.5 isolates is ~0.27 SNPs per genome per year, which is approximately 3-fold less than previous *S.* Typhi studies focusing on the rapidly spreading multiple drug resistant H58/genotype 4.3.1 (13, 85). These results may indicate different environmental pressures in Samoa or an intrinsically more slowly evolving *S.* Typhi population in a geographically remote island setting. Additionally, it is remarkable that while there have been sporadic isolations of non-3.5.4/3.5.3 in Samoa since 1983, and most likely before, there has not been the replacement of the 3.5.4/3.5.3 genotypes by these incoming external genotypes. This suggests that there may be other factors, such as host genetics (94, 95), microbiome (96), or environmental factors (97), as well as unknown mechanisms of transmission, that support the maintenance of the 3.5.4/3.5.3 genotypes in Samoa and their endurance, heretofore, despite classical typhoid control efforts.

We have assessed Samoan *S.* Typhi for intrinsic factors that may provide such a competitive or survival advantage. While many putative protein-coding sequences missed through mapping analyses were uniquely present in the Samoan *S.* Typhi isolates and absent from the diverse set of reference genomes as well as vice-versa, the functions of most of these genes are currently unknown. An ~106-kb phenotypically-cryptic pHCM2 molecule that does not encode resistance or virulence factors (98) was detected in Samoan *S.* Typhi isolates as early as 1994 and has retained identical plasmid length and gene content 25 years later (Fig. 4B). The proportion of Samoan *S.* Typhi bearing IncFIB(pHCM2) was more than double the observed proportion of pHCM2-containing isolates in the global collection. This significant (p<0.01) enrichment of this conserved plasmid in the Samoan *S.* Typhi population may be due to enhanced plasmid acquisition and stability, an intrinsic advantage for its maintenance, or an artifact of the local *S.* Typhi population. Our genomic content analyses of pHCM2 from Samoa, India, Bangladesh, and Vietnam (Fig. 4B) revealed numerous genes encoding hypothetical proteins, and a limited number of proteins associated with gene regulation, central metabolism or mobile elements, but none are predicted to impart an advantage based on the current functional annotations (Table S7). This remarkable gene conservation across significant geographic distances and time suggests that some functions beneficial to their persistence or to the *S.* Typhi host may be encoded on this plasmid, but these are yet to be elucidated.

Finally, we observed that there has been limited development of AMR in Samoa, as mutations associated with AMR were detected in only 15/306 (4.9%) of Samoan isolates (Fig. 2, Table S1-S2). Pan-susceptibility to first-line antibiotics was identified in 2018 to 2020 by the clinical microbiology laboratories in Samoa (Table S1) and our examination of genomic features associated with antibiotic resistance largely corroborates this observation. There was incongruence between genotype and phenotype observed; however, the reason for this incongruence is presently unknown. This finding should give pause to assigning phenotypic resistance and to making clinical or regulatory decisions based solely on WGS data. A limited study of antibiotic prescriptions and usage in 2004 in Samoa revealed higher levels of prescribing (66.4% percent of prescriptions included an antibiotic) and a greater reliance on penicillins (63% of defined daily doses per 1000 inhabitants per day) compared with other low- and middle-income countries (99). Unmeasured over-the-counter sales of antibiotics were also reported (99). There appears to be less AMR development among the endemic Samoan strains of *S.* Typhi when compared to other countries in spite of liberal antibiotic usage. It is currently unclear if this is associated with the dominant and stable 3.5.4/3.5.3 genotypes or the patterns of antibiotic usage.

Limitations of this study include inconsistent sampling over time, which is an issue with many genomic studies in resource-restricted locations. Therefore, we relied on historical travel-associated isolates collected in Australia and New Zealand where detailed epidemiologic data was not always available. For example, singular historical isolations of genotypes 2.2.1, 2.3.2, and 3.5 in 2012, 1983, and 1985, respectively, were detected in individuals in New Zealand and Australia who had recent travel history to Samoa. These could indicate under-sampled genotypes that were once common in Samoa. However, these genotypes are identified globally and could have been mischaracterized as Samoa-associated importations if other travel linkages were not disclosed. Since 2018, the Samoa TFCP has paired WGS with improved blood culture surveillance and household investigations to mitigate this limitation and provide contiguous years of genome sequencing. Future investment in local sequencing capacity and implementation of rapid bioinformatics pipelines of public health utility would greatly strengthen local and regional infectious disease surveillance with complementary WGS data.

In sum, we have determined that typhoid fever in Samoa is caused by a Samoa-restricted *S.* Typhi population comprised of rare but stable genotypes 3.5.4 and 3.5.3 that have not developed antimicrobial resistance over decades of endemicity and suboptimal antibiotic prescribing practices. However, this relatively simple situation, where first-line drugs remain effective in treating typhoid fever in Samoa, could suddenly change. *S.* Typhi belonging to H58/genotype 4.3.1 encoding drug resistance, as well as enhanced virulence (8), have emerged and spread internationally (24, 25, 100). While genotype 4.3.1 was not detected in this collection of Samoan *S.* Typhi, Samoa has endured persistently endemic typhoid fever for decades and could be vulnerable to the development of antimicrobial resistance in an endemic isolate or the importation and dissemination of a resistant clone similar to H58/genotype 4.3.1

Through this study, we have established a Samoa-specific *S.* Typhi genomic framework that will allow for enhanced typhoid surveillance. For example, pairing these WGS data with traditional epidemiology and geospatial clustering analyses can provide insights into the transmission mechanics of *S.* Typhi in Samoa, which may be applicable to other islands in Oceania. Such data can inform the setting of interventions to be carried out by the Samoa MoH and can help monitor their progress, such as assessing the *S.* Typhi genomic population structure before and after mass vaccination with Vi conjugate vaccine, which commenced in Samoa in August 2021 (101). Should AMR *S*. Typhi emerge in Samoa, timely WGS analysis can likely provide a genomic examination of the phenotypic AMR, which may implicate the geographic origin (locally or internationally). Finally, once the incidence of acute cases is drastically reduced in high burden age groups following the current mass vaccination program, WGS data can assist the MoH during the Consolidation Phase of the Samoa TFCP, which is intended to actively seek out silent chronic typhoid carriers who will constitute the long-term human reservoir of *S*. Typhi.

## ACKNOWLEDGEMENTS

The authors wish to acknowledge the staff at the Ministry of Health of Samoa for their administrative and technical support of the Samoa Typhoid Fever Control Program, the staff at the Microbiological Diagnostic Unit Public Health Laboratory, Jane Han for her logistical support of the Samoa Typhoid Fever Surveillance Initiative, and Corin Yeats for his technical support with Pathogenwatch.

This work was supported by the Bill & Melinda Gates Foundation OPP1194582 (INV-000049) (PI: Prof. Myron M. Levine). Under the grant conditions of the Foundation, a Creative Commons Attribution 4.0 Generic License has already been assigned to the Author Accepted Manuscript version that might arise from this submission. M. J. S. received research support in part by federal funds from National Institutes of Health under National Institute of Allergy and Infectious Diseases grants F30AI156973 (PI: Michael J. Sikorski) and U19AI110820 (PI: David A. Rasko), as well as National Institute of Diabetes and Digestive and Kidney Diseases training grant T32DK067872 (PI: Jean-Pierre Raufman). M. M. L. is supported in part by the Simon and Bessie Grollman Distinguished Professorship at the University of Maryland School of Medicine. The funders had no role in the design and conduct of the study; collection, management, analysis, and interpretation of the data; preparation, review, or approval of the manuscript; and decision to submit the manuscript for publication.

M. M. L. reports grants from Bill & Melinda Gates Foundation during the conduct of the study; in addition, M. M. L. has a patent entitled “Broad spectrum vaccine against typhoidal and nontyphoidal *Salmonella* disease” (US 9,011,871 B2) issued. R. M. R.-B. reports grants from Bill & Melinda Gates Foundation, and non-financial support from the Government of Samoa during the conduct of the study. All other authors report no potential conflicts.

